# Effective connectivity of the hippocampus can differentiate patients with schizophrenia from healthy controls: a spectral DCM approach

**DOI:** 10.1101/2020.01.12.20017293

**Authors:** Lavinia Carmen Uscătescu, Lisa Kronbichler, Renate Stelzig-Schöler, Brandy-Gale Pearce, Sarah Said-Yürekli, Luise Antonia Reich, Stefanie Weber, Wolfgang Aichhorn, Martin Kronbichler

## Abstract

We applied spectral dynamic causal modelling (spDCM; Friston et al., 2014) to analyze the effective connectivity differences between the nodes of three resting state networks (i.e. Default mode network/DMN, Salience network/SAN and Dorsal attention network/DAN) in a dataset of 31 healthy controls (HC) and 25 patients with schizophrenia (SZ), all male. Patients showed increased connectivity from the left hippocampus (LHC) to the dorsal anterior cingulate cortex (DACC), right anterior insula (RAI), left frontal eye fields (LFEF) and the bilateral inferior parietal sulcus (LIPS & RIPS), as well as increased connectivity from the right hippocampus (RHC) to the bilateral anterior insula (LAI & RAI), right frontal eye fields (RFEF) and RIPS. Moreover, negative symptoms predicted the connectivity strengths from the LHC to the DACC, the left inferior parietal sulcus (LIPAR) and the RHC, while positive symptoms predicted the connectivity strengths from the LHC to the LIPAR and from the RHC to the LHC. These results reinforce the crucial role of hippocampus dysconnectivity in SZ pathology and its potential as a biomarker of disease severity.

## 1. Introduction

Schizophrenia is a common and debilitating psychiatric disorder, diagnosed in about 1% of the world’s population (Bhugra, 2005). It is characterized by negative (e.g. disorganized thoughts and language, attention and memory deficits) and positive (e.g. hallucinations and delusions) symptoms. Assessing potential brain markers of SZ onset, progression and severity has consequently led to increased research interest and to the development of various data acquisition and analysis methods.

Resting state functional magnetic resonance imaging (rsfMRI) is a particularly convenient acquisition modality given its short duration and lack of cognitive demands, which makes it particularly suitable for studies involving patients. From the acquired rsfMRI time-series one can identify resting state networks (RSNs; Greicius et al., 2003), which are patterns of co-activated brain areas that share a common function (e.g. Greicius, 2008). Some RSNs, like the Default mode network (DMN), increase their activity during rest and internally directed cognitive activity (Raichle, 2015). The nodes belonging to the so-called “core” DMN are the posterior cingulate cortex (PCC), the medial prefrontal cortex (MPFC), and the left and right inferior parietal cortex (LIPAR; RIPAR); in addition, the left and right hippocampi (LHC; RHC) are sometimes included (e.g. Ushakov et al., 2016), thus forming an “extended” DMN. Other networks, such as the Salience network (SAN) and the Dorsal attention network (DAN) show increased activity during externally directed cognitive processes (Uddin, 2016; Vossel, Geng & Fink, 2014). The SAN comprises the dorsal anterior cingulate cortex (DACC), and the bilateral anterior insula (LAI; RAI). The DAN is composed of the bilateral frontal eye fields (LFEF; RFEF), and the bilateral inferior parietal sulcus (LIPS; RIPS).

The study of RSNs opened new possibilities for understanding SZ pathology. Directed connectivity via spectral dynamic causal modelling (spDCM) is the most recent approach dedicated to the analysis of rsfMRI data (Friston et al., 2014). Compared to its stochastic counterpart, spDCM is more computationally efficient, and seems especially promising for clinical research as it has proven to be more sensitive to group differences (Razi et al., 2015). So far, only few spDCM studies involving SZ patients have been published (Chahine et al., 2016; Cui et al., 2015; Fang et al., 2018; Graña, Ozaeta and Chyzhyk, 2017a, 2017b), and only one assessing hippocampal dysconnectivity (Cui et al., 2015). These authors found, in a sample of first episode SZ, increased connectivity from the left dorsolateral prefrontal cortex to the LHC, but decreased connectivity from the right anterior cingulate cortex to the RHC and from the LHC to the dorsolateral prefrontal cortex. Previous stochastic DCM research in SZ samples also showed that the hippocampus is differentially connected to other brain areas during rest. For example, Li et al. (2017) found significantly decreased connectivity from the auditory cortex to the hippocampus in patients with auditory verbal hallucinations compared to HC. Additionally, Lefebvre et al., (2016) further provided evidence supporting the involvement of the connectivity from the hippocampus to the SAN as being linked to the onset of hallucinations. These findings therefore promote the notion that the hippocampus might be a core driving area of hallucinatory experiences, likely via hyperdopaminergic mechanisms (Kapur, 2003; Lodge and Grace, 2011; Winton-Brown et al., 2014; Modinos et al., 2015).

Research using DCM has further shown that the hippocampus connectivity is not only altered in SZ patients, but also in unaffected 1^st^ degree patients’ relatives and in persons at ultra-high risk (UHR) for psychosis, thus appearing to hold a promise as a biomarker of psychosis proneness. For example, Xi et al. (2016) showed, using stochastic DCM, that unaffected 1^st^ degree relatives of SZ, compared to HC, displayed increased connectivity from the left anterior cingulate cortex to the RHC, but decreased connectivity from the right anterior cingulate cortex to the RHC. In a task-based fMRI DCM study, Winton-Brown et al. (2017) further showed that the dysconnectivity of the hippocampal–basal ganglia–midbrain network during reward, novelty and aversion processing can differentiate UHR from HC.

The hippocampus, as a hub for memory processes (Battaglia et al., 2011; Bernal-Casas et al., 2013) and for the whole connectome (Mišić et al., 2014), can offer additional clues regarding the mechanism of cognitive dysfunctions in SZ. In a task-based DCM study investigating associative learning, Banyai et al. (2011) found increased intrinsic connectivity in SZ compared to HC from the prefrontal cortex to the hippocampus and from the hippocampus to the inferior temporal and superior parietal, even after controlling for the learning rate. In a memory task-based DCM study, Benetti et al., (2009) showed increased connectivity from the right posterior hippocampus to the right inferior frontal gyrus in HC compared to both first episode SZ and UHR, although the latter two groups did not differ significantly. Behaviourally, SZ, but not UHR, showed impaired memory performance compared to HC. Taken together, these results support the notion that the dysfunctional prefrontal-hippocampal coupling underlies memory deficiencies in SZ, and, moreover, that it could be used as a reliable intermediate phenotype biomarker (Bächner and Meyer-Lindenberg, 2017).

In the present study, we employed spDCM to investigate the effective connectivity differences between SZ and HC. First, we looked at the resting state effective connectivity between individual nodes belonging to the DMN, the SAN and the DAN, and the relationship between symptom severity and connection strengths. Second, we analyzed group differences in the hierarchical structure of these three networks, similar to Nee & D’Esposito (2016) and Zhou et al. (2018a). Third, using voxel-based morphometry (VBM) we analyzed gray matter volume (GMV) group differences in all the nodes included in our spDCM and re-assessed group differences in connection strengths while controlling for GMV.

## 2. Materials and Methods

### 2.1. Participants

In the current study, 25 male patients with an official diagnosis of schizophrenia and 31 age and education matched male healthy controls were included. The patients were recruited through the Department of Psychiatry, Psychotherapy and Psychosomatics in Salzburg, Austria. At the time of scanning, patients were medicated and clinically stable. Symptom severity, as assessed with PANSS (Kay, Fiszbein and Opler, 1987), was found to be mild. Two of the patients did not complete the PANSS assessment but did complete the resting state scanning session. To ensure that controls were indeed free from any mental disorders, they were screened using the Mini-International Neuropsychiatric Interview (Sheehan et al., 1998). More details about the recruitment and assessment of the participants included in the present study can be found in Kronbichler et al. (2018). Demographic data and clinical scores are presented in Table 1.

**Table 1:** Demographic data of patients with schizophrenia (SZ) and healthy controls (HC). Two participants from the SZ group did not complete the Positive and Negative Syndrome Scale (PANSS) assessment. PANSS+ reflects the severity of positive symptoms, while PANSS-that of negative symptoms. PANSS g reflects the general psychopathology index.

| Group | n | Age | Illness duration<br>(years)<br>(n = 24) | PANSS+<br>(n = 23) | PANSS-<br>(n = 23) | PANSS g<br>(n = 23) |
| --- | --- | --- | --- | --- | --- | --- |
| SZ | 25 | 26.52 ± 5.03 | 4 ± 4.96 | 14 ± 5.72 | 15.48 ± 7.02 | 1.65 ± 0.94 |
| HC | 31 | 25.43 ± 4.3 |  |  |  |  |

### 2.2. Data Acquisition and Preprocessing

Imaging data were acquired with a Siemens Magnetom Trio 3 T scanner (Siemens AG, Erlangen, Germany) using a 32-channel head coil. Functional images were obtained with a T2*-weighted gradient echo EPI sequence (TR 2,250 ms, TE 30 ms, matrix 64 mm × 64 mm, FOV 192 mm, flip angle 70°). Additionally, a gradient echo field map (TR 488 ms, TE 1 = 4.49 ms, TE 2 = 6.95 ms) and a high-resolution (1 mm × 1 mm × 1 mm) structural scan with a T1-weighted MPRAGE sequence were recorded from each participant. A total of 110 resting state volumes (TR 2.250 ms, TE 1 = 4.49 ms, TE 2 = 6.95 ms) were additionally acquired. During the resting state sequence, participants were instructed to keep their eyes open and look at a fixation cross on the screen, while letting their mind wander.

Preprocessing of resting state data was performed using SPM12; functional scans were realigned, de-spiked, unwarped, corrected for geometric distortions, and slice time corrected.

They were also normalized to MNI space and co-registered to the corresponding skull-stripped structural images, and afterwards resampled to 3 mm × 3 mm × 3 mm voxels and smoothed with a 6 mm FWHM Gaussian kernel. Preprocessing steps of structural data from this sample have previously been reported elsewhere (Kronbichler et al., 2018).

### 2.3. Data Analysis

The three RSNs of interest to our current study were identified via spatial, constrained ICA, as implemented in the Group ICA for fMRI Toolbox (GIFT, http://mialab.mrn.org/software/gift) (Calhoun et al. 2001). The spatial templates used to constrain the ICA were those of Shirer et al. (2012), downloaded from http://findlab.stanford.edu/research. Nodes coordinates at the group-level are summarized in Table 2. The layout of the three networks, as mapped onto our sample data, can be seen in Figure 1.

**Table 2:**
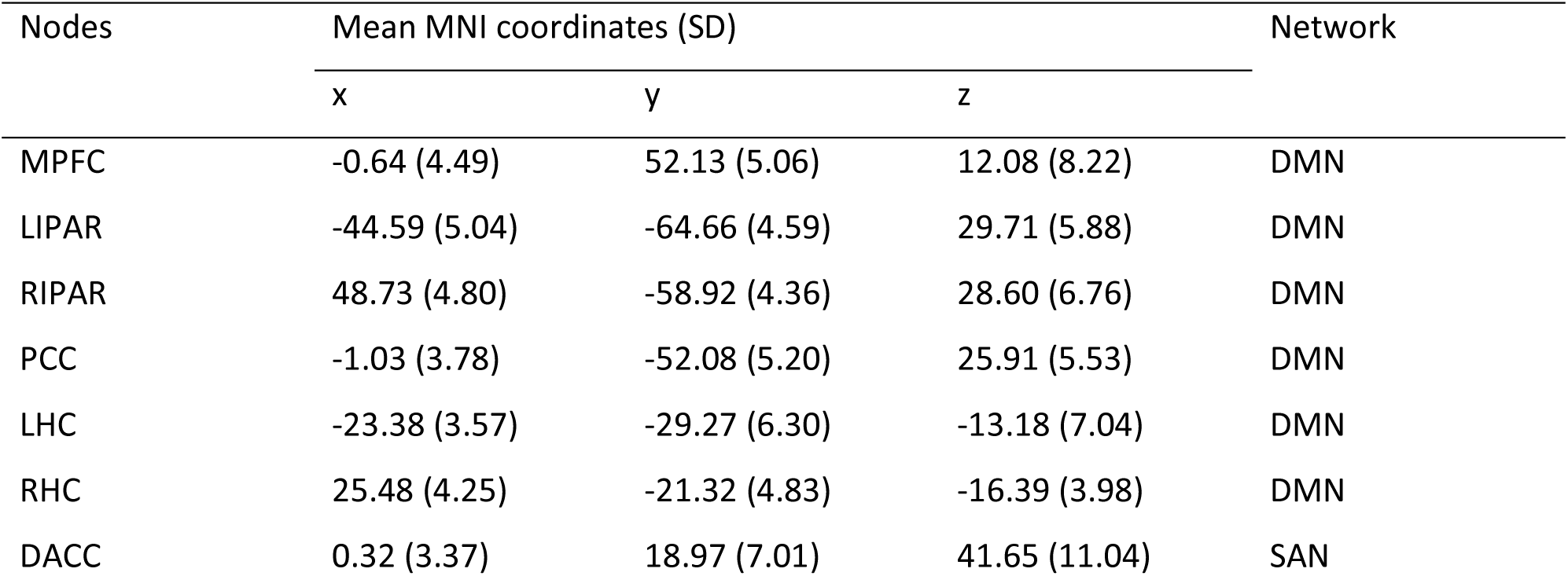
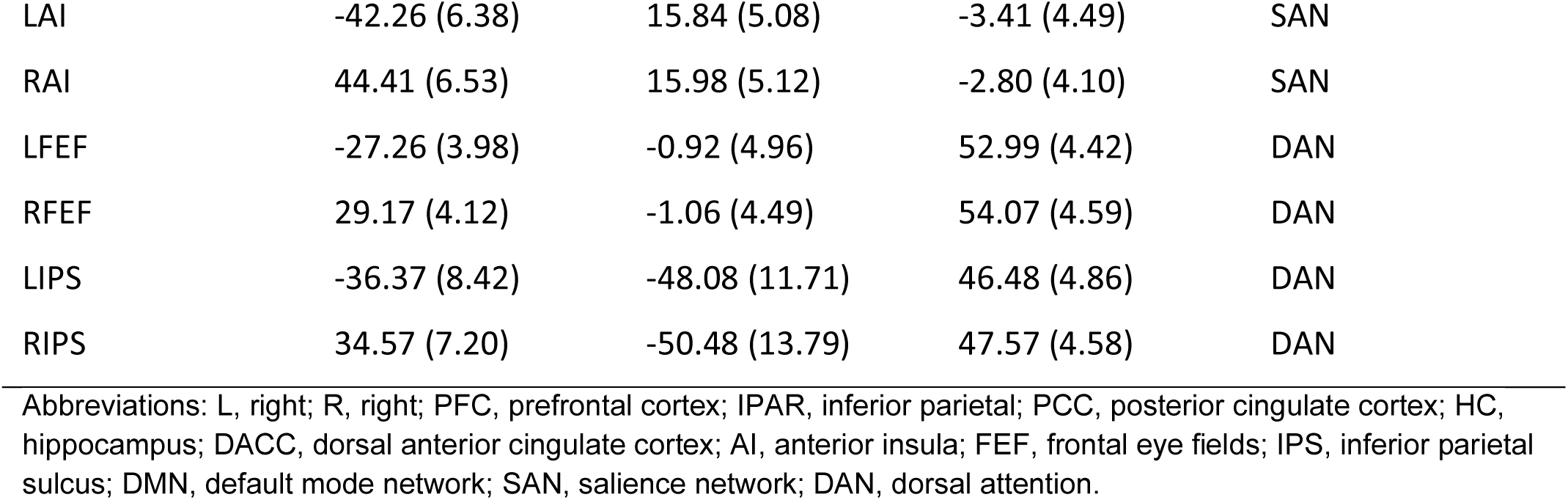
Group-level coordinates of individual nodes. Standard deviations are given in parentheses.

| Nodes | Mean MNI coordinates (SD) |  |  | Network |
| --- | --- | --- | --- | --- |
|  | x | y | z |  |
| MPFC | -0.64 (4.49) | 52.13 (5.06) | 12.08 (8.22) | DMN |
| LIPAR | -44.59 (5.04) | -64.66 (4.59) | 29.71 (5.88) | DMN |
| RIPAR | 48.73 (4.80) | -58.92 (4.36) | 28.60 (6.76) | DMN |
| PCC | -1.03 (3.78) | -52.08 (5.20) | 25.91 (5.53) | DMN |
| LHC | -23.38 (3.57) | -29.27 (6.30) | -13.18 (7.04) | DMN |
| RHC | 25.48 (4.25) | -21.32 (4.83) | -16.39 (3.98) | DMN |
| DACC | 0.32 (3.37) | 18.97 (7.01) | 41.65 (11.04) | SAN |
| LAI | -42.26 (6.38) | 15.84 (5.08) | -3.41 (4.49) | SAN |
| RAI | 44.41 (6.53) | 15.98 (5.12) | -2.80 (4.10) | SAN |
| LFEF | -27.26 (3.98) | -0.92 (4.96) | 52.99 (4.42) | DAN |
| RFEF | 29.17 (4.12) | -1.06 (4.49) | 54.07 (4.59) | DAN |
| LIPS | -36.37 (8.42) | -48.08 (11.71) | 46.48 (4.86) | DAN |
| RIPS | 34.57 (7.20) | -50.48 (13.79) | 47.57 (4.58) | DAN |

Abbreviations: L, left; R, right; PFC, prefrontal cortex; IPAR, inferior parietal; PCC, posterior cingulate cortex; HC, hippocampus; DACC, dorsal anterior cingulate cortex; AI, anterior insula; FEF, frontal eye fields; IPS, inferior parietal sulcus; DMN, default mode network; SAN, salience network; DAN, dorsal attention.

**Figure 1:**
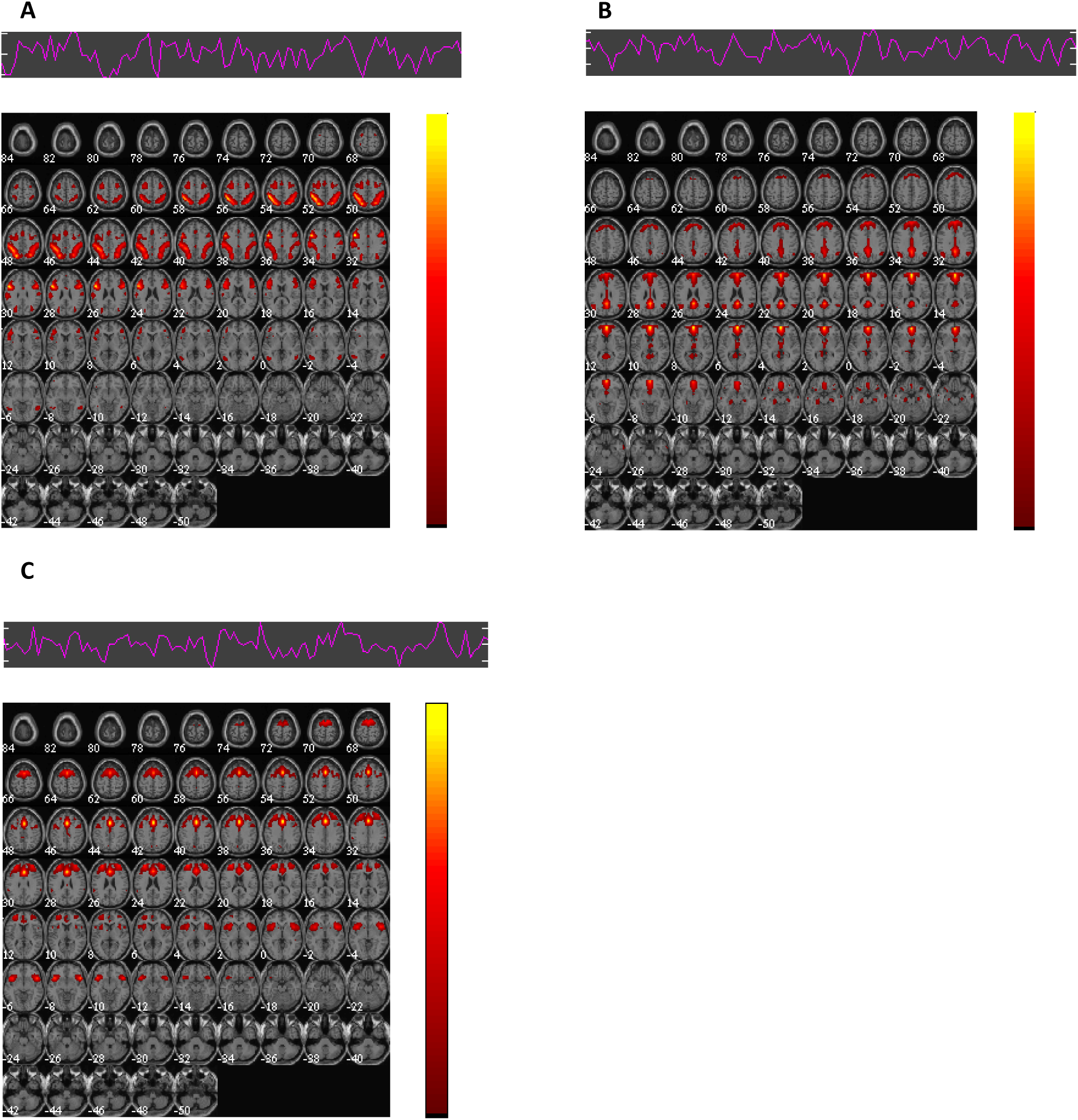
The three RSNs we used in our spDCM analysis, as illustrated at the group level in our entire sample: **A**, Dorsal attention network (DAN); **B**, Default mode network (DMN); **C**, Salience network (SAN). No significant group differences between the time series of these components were found between HC and SZ (FWE corrected at p < .05).

To obtain individual time-courses for each node belonging to the three RSNs, the following steps were performed, following the identification of the ICA components corresponding to the DMN, DAN and SAN. First, group masks were created for each RSN, based on one sample t tests of the respective component. This was followed by a 1^st^ level design using the AROMA motion corrected preprocessed resting state files (Pruim et al., 2015), the single subject reconstructed time-courses for our three independent components, as well as covariates reflecting 5 principal components (PCs) of average signal for a WM mask (based on each subject’s WM mask) and 5 PCs for CSF mask (based on each subject’s ventricle mask) plus the 6 realignment parameters. Finally, individual time-courses for the DCM analysis were extracted from these 1^st^ level designs by selecting the subjects’ local maximum from each ROI (constrained by the group mask for the respective ROI) for the contrast testing the association between the subject’s BOLD signal and the subjects ICA time-course. These local maxima were extracted as an 8 mm radius centered sphere.

Spectral DCM is a deterministic, linear alternative to stochastic DCM. Because the spectral generative model relies on second order statistics (i.e., cross spectra) of original timeseries, estimating their varying hidden states can be circumvented by estimating their time invariant covariance. This renders the model more computationally efficient, since the inversion scheme now only requires estimating the model’s parameters and hyperparameters. A detailed mathematical treatment of spDCM can be found in Friston et al. (2014) and Razi et al. (2015).

The spDCM analysis was performed in SPM12 r7487 (Wellcome Trust Centre for Neuroimaging, London, UK; code available at: https://github.com/spm/spm12). At the first level, fully-connected models (i.e. between all nodes plus self-inhibitory connections) were estimated for each subject individually. At the second level analysis, the group model was built using the fully connected models from each subject, therefore capturing the between-subjects effect on each of the modelled connections. Group differences in connection strength between the pre-defined nodes were assessed using a parametric empirical Bayes (PEB) model (Friston, Zeidman and Litvak, 2015; Friston et al., 2016). The PEB is a hierarchical approach, in which the posterior density of model parameters is constrained by that of the previous level. Finally, post-hoc search was performed, which identifies and eliminates those parameters which do not contribute to model evidence.

Voxel based morphometry (VBM) was performed using the REX toolbox (available at https://web.mit.edu/swg/software.htm). Gray matter volume (GMV) was extracted from each participant for each of the nodes used in our spDCM analysis. All the other statistical analyses were performed in R 5.2 (R Core Team, 2018).

## 3. Results

### 3.1. Effective/directed connectivity

The connections with a posterior probability of more than 95% are depicted in Figure 2 below. To ease comprehension of results, these connections are further mapped onto a brain template in Figures 3 and 4.

**Figure 2:**
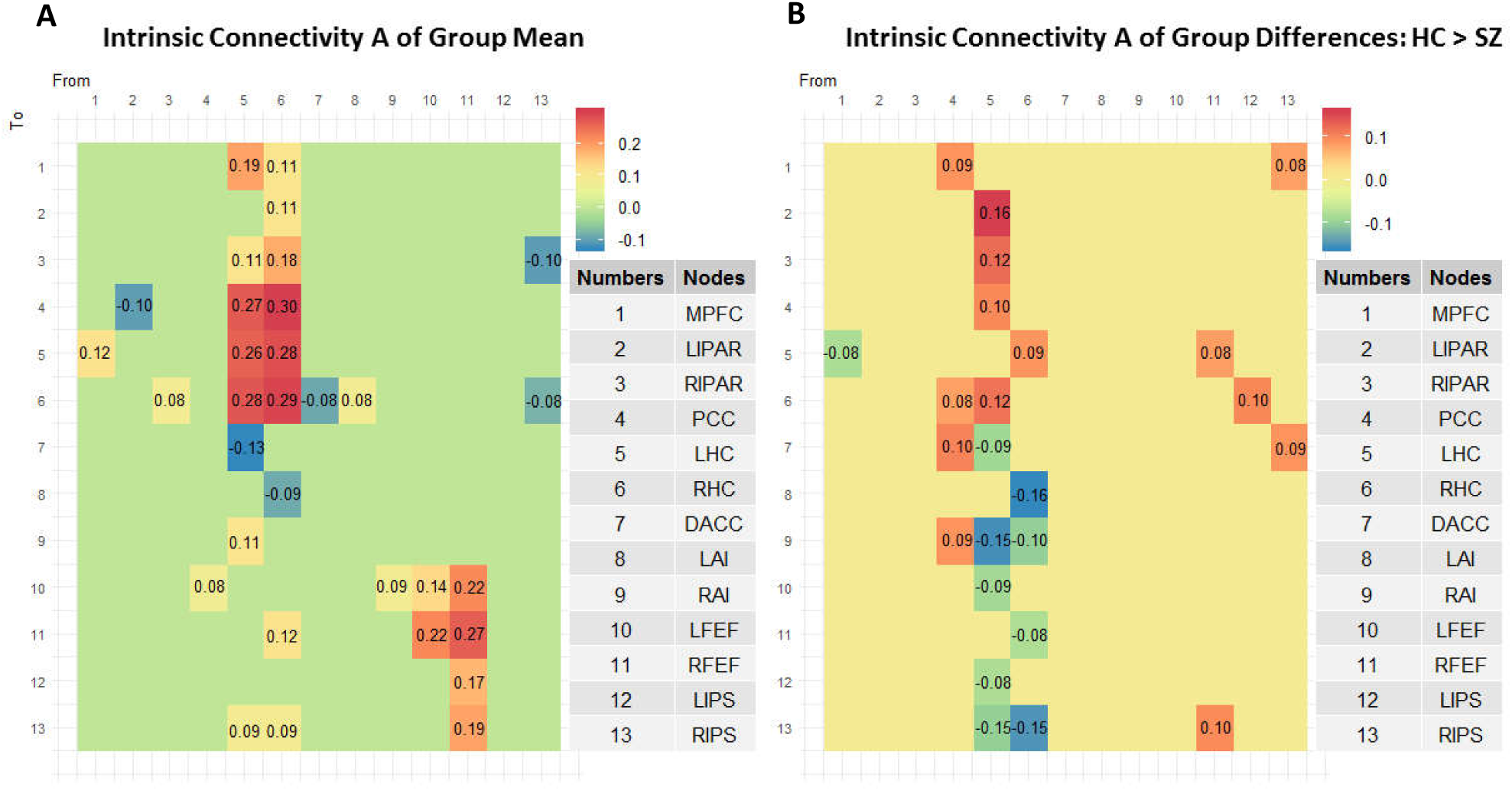
**A**, Intrinsic connectivity matrix reflecting group means in effective/directed connectivity between the 13 nodes. Only connections with a posterior probability > 0 .95 are displayed; **B**, Intrinsic connectivity matrix A reflecting group differences in effective/directed connectivity between the 13 nodes. Only connections with a posterior probability > 0 .95 are displayed. The results reflect connection strengths as a difference between those in the HC group and those in the SZ group (I.e. HC > SZ); the colour gradient therefore reflects positive values for those connections which were stronger in HC than in SZ, and negative values for those connections which were stronger in SZ compared to HC.

**Figure 3:**
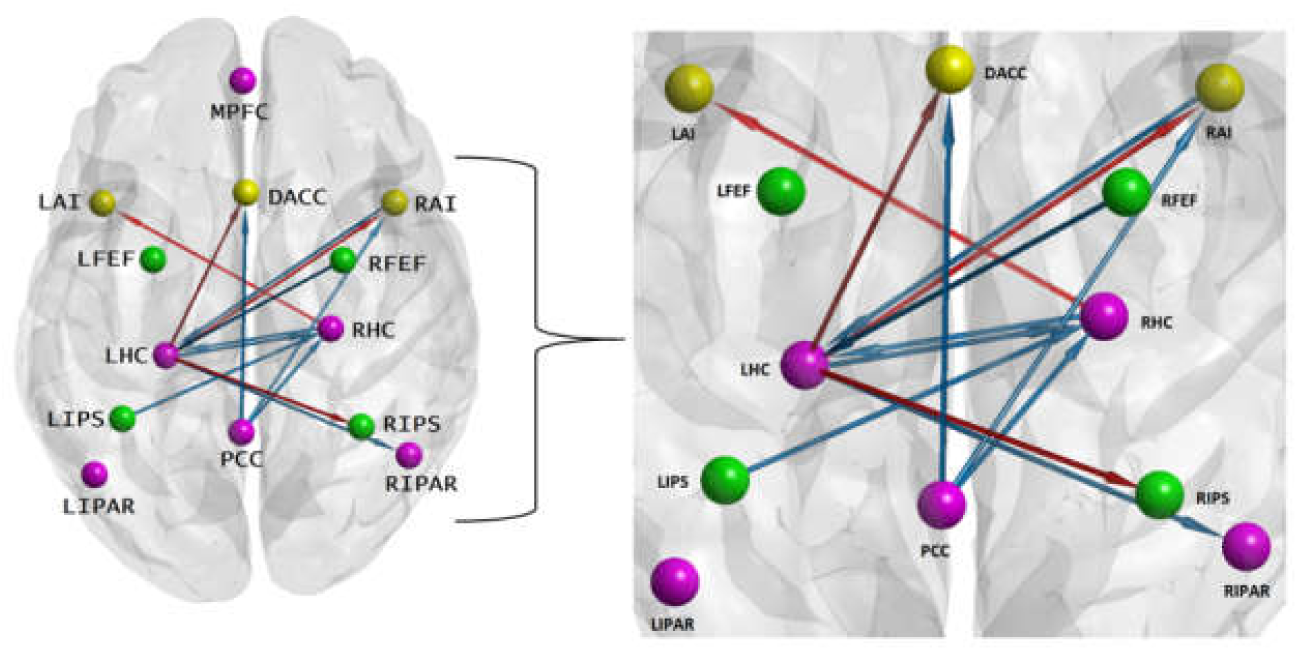
Inter-hemispheric directed connections. Legend: orange arrows, SZ; blue arrows, HC; magenta nodes, DMN; green nodes, DAN; yellow nodes, SAN. BrainNet Viewer software (Xia, Wang & He, 2013; http://www.nitrc.org/projects/bnv/) was used to map the directed connections onto the brain surface.

**Figure 4:**
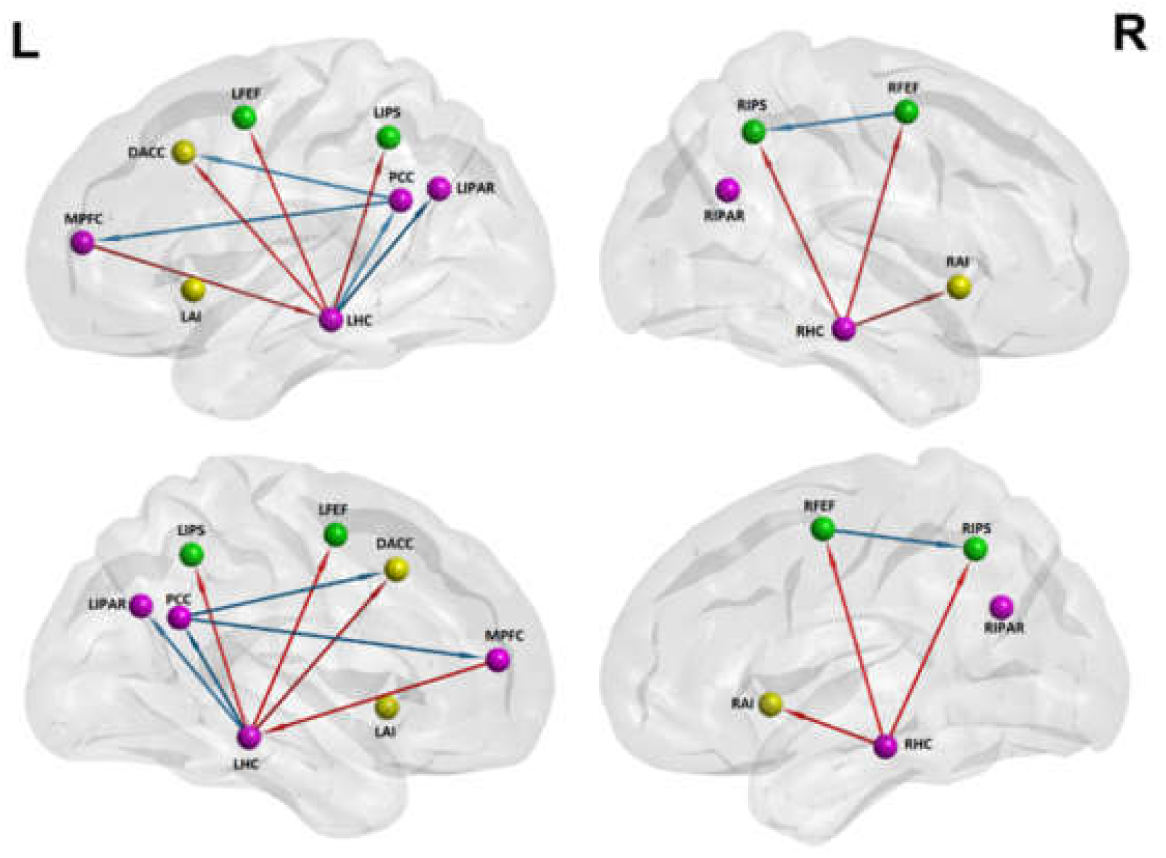
Intra-hemispheric directed connections. Legend: orange arrows, SZ; blue arrows, HC; magenta nodes, DMN; green nodes, DAN; yellow nodes, SAN. BrainNet Viewer software (Xia, Wang & He, 2013; http://www.nitrc.org/projects/bnv/) was used to map the directed connections onto the brain surface.

Upon visual inspection, it appears that the LHC and RHC drive most of the significant connections, both at the whole group level and when looking at group differences. We modelled each column (i.e. out-going connections) from our connectivity matrices (Fig. A and B) as a binomial distribution with 13 possible outcomes (as there are 13 potential connections stemming from each node, including the self-connections). For each of the 13 possible outcomes, there is an equal likelihood of there being a connection present (so a 0.5 probability for a connection being present or absent). Knowing what the number of connections stemming from each node is, we obtain that at the chance level, each node from our connectivity matrix can drive connections into 6 other nodes. Therefore, any number of connections strictly lower than 6 would fall below chance level, while any number strictly higher than 6 and up to (and including) 13 would be above chance level. We can thus see that at the whole group level, it is the LHC and the RHC who drive an above chance level number of connections, with the LHC remaining dominant also when comparing HC to SZ. A mathematical treatment of the algorithm used for this analysis can be found in Loader (2002).

To additionally quantify the hierarchical connection strength of our three RSNs, we computed, from the (unthresholded) mean group level connection strengths (illustrated in Figure 1.A), average connectivity strengths of each of our three RSNs, as well as between any give pair, bidirectionally (see Figure 5 below). To achieve this, we followed the procedures described by Nee & D’Esposito (2016) and Zhou et al. (2018a). Additionally, like Zhou et al. (2018a), we also computed the uncertainty of each between-network connection strength, which we report in parentheses, as standard deviations (see Figure 5 below). In the top left panel of Figure 5, we illustrate hierarchical strengths computed on the mean connectivity strengths of the entire group mean (comprising both HC and SZ). The top right panel of Figure 5 shows the hierarchical strengths computed on the group difference connectivity strengths of (HC - SZ). In the bottom left panel of Figure 5, the hierarchical strengths for HC are illustrated, while the hierarchical strengths for SZ are illustrated on the bottom right panel of Figure 5.

**Figure 5.**
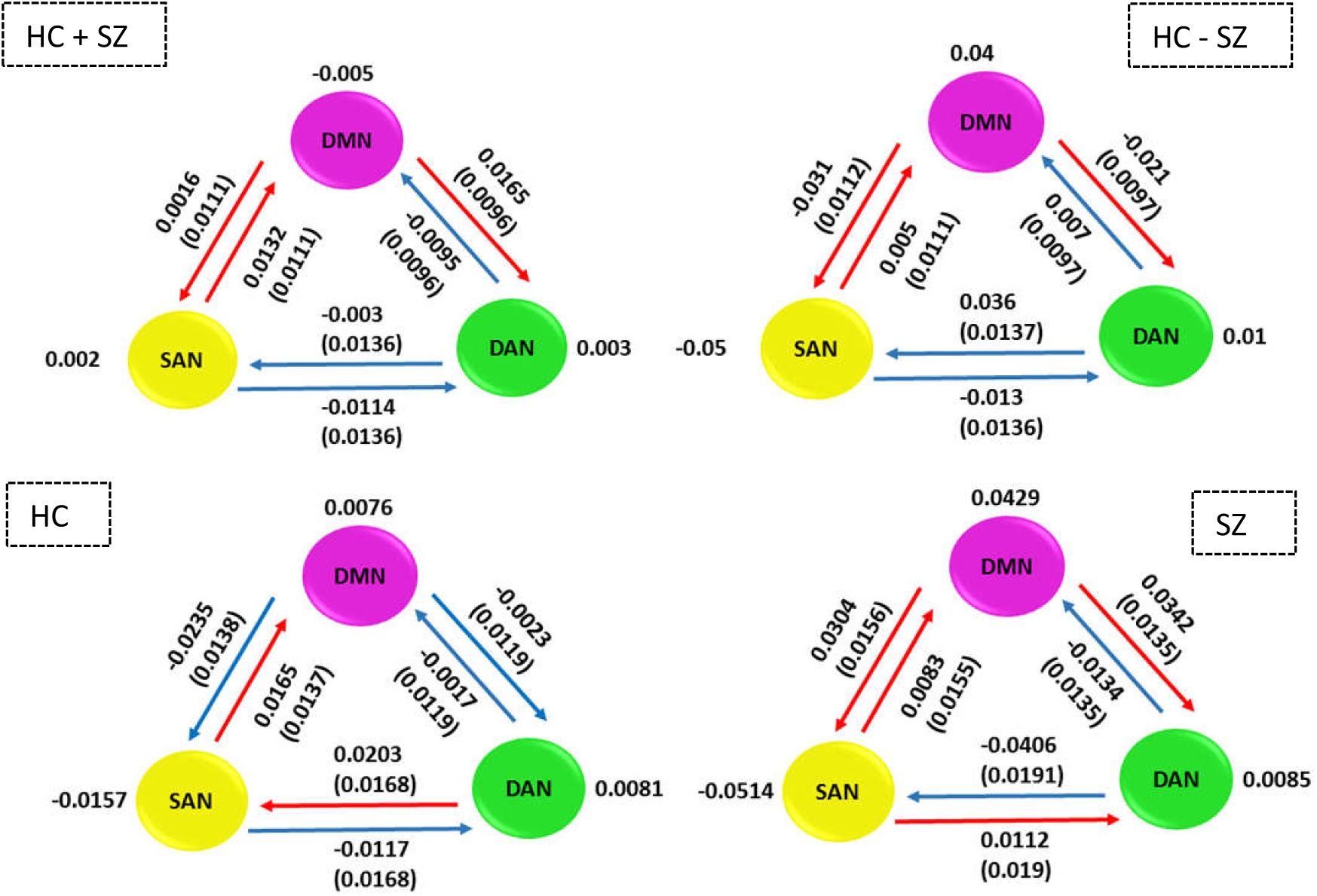
Hierarchical organization between the three RSNs, as well as individual hierarchical strengths. The uncertainty is reported in parentheses as standard deviation about each hierarchical connection. Inhibitory connections are rendered with blue arrows, while the excitatory ones with red arrows. The top left graph illustrates the hierarchical organization for the mean connectivity of entire group (HC + SZ), while the top right one shows the hierarchical strengths for group differences (HC – SZ). The bottom left panel shows hierarchical strength for HC only, while the bottom right one for SZ only.

Note that in all three situations the SAN excites the DMN, while the DAN inhibits the DMN. While the second result is in accordance with what Zhou et al. (2018a) reported, in our dataset, the SAN excites the DMN for both HC and SZ. One potential explanation for this discrepancy could reside in the fact that these authors only considered the core DMN areas, while we additionally incorporated the hippocampi. An interesting finding in our current research is that in HC it is the DAN which excites the SAN, whereas in SZ, it is the other way around. Additionally, in HC, the DMN inhibits the DAN, but in SZ, it excites the DAN.

### 3.2. The relation between symptom severity and the strength of directed connectivity

We assessed the relation between individual connectivity parameter strength and symptom severity (as reflected by the PANSS scores) from the SZ group. Because in some cases we were concerned about the possibility of outlier points, but could not substantiate a decision to remove these, we decided to fit both simple linear and robust regression models. Positive and negative symptoms significantly predicted some of the directed connection strengths, but not the general psychopathology index. Details are given below.

#### Negative Symptoms Severity and Connectivity Strength

A negative correlation *r* = − .53 between the severity of negative symptoms and connectivity strength from the LHC to the DACC was found, and a simple linear regression model (see Figure 6. A) further showed a significant (*p* = .009) relation between the two variables. The slope coefficient for the severity of negative symptoms was −0.057, so connectivity strength of the parameter decreases by 0.057 for every additional one unit increase of negative symptom severity. The adjusted R^2^ value was 0.2444, which indicates that 24.44% of the variation in LHC to DACC connectivity strength can be explained by negative symptom severity.

**Figure 6.**
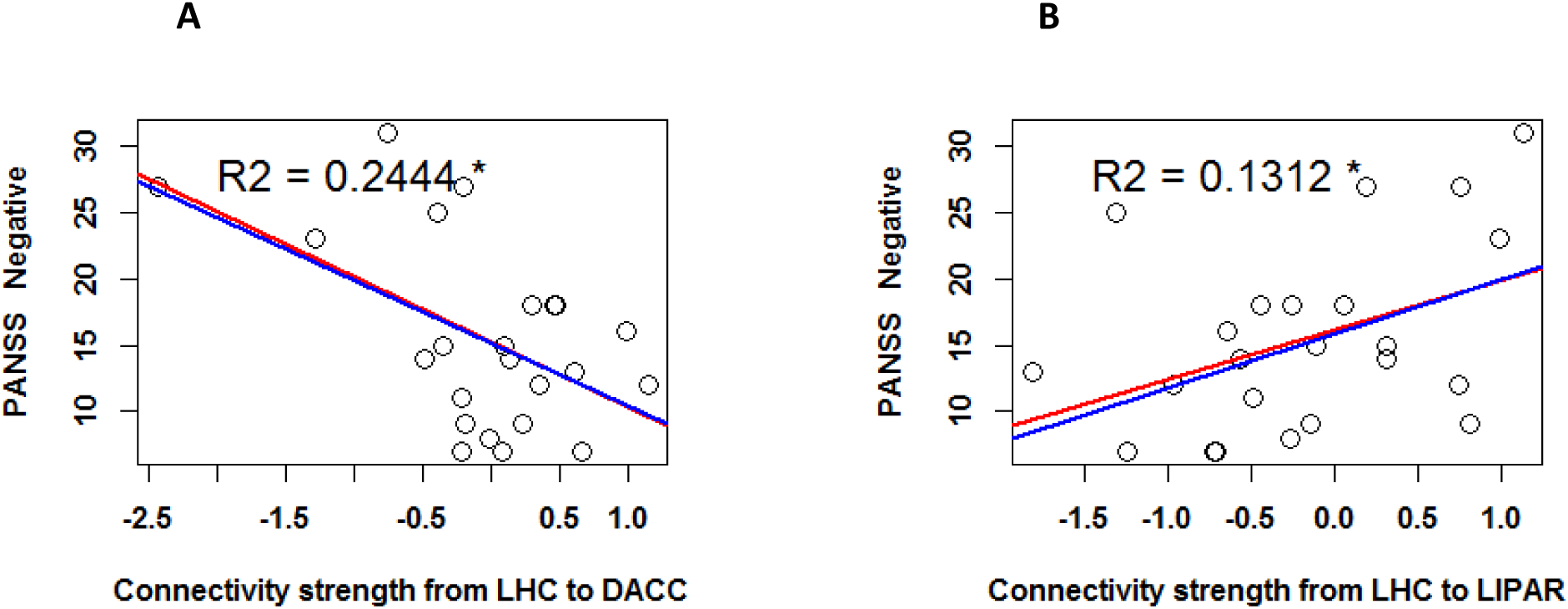
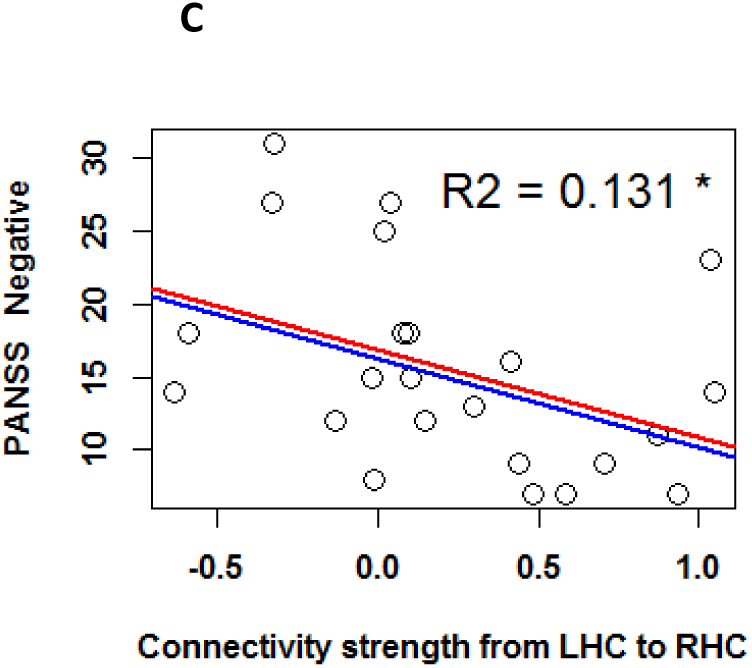
Linear regression with severity of negative symptoms (as reflected by PANSS scores) as regressor. The predicted variables are the connectivity strengths of the following parameters: **A**: from the LHC to the DACC **B**: from the LHC to the LIPAR, and **C**: from the LHC to the RHC. The red regression line reflects the fit of the simple linear regression model, while the blue line reflects that of the robust regression model. The plotted R^2^ values reflect the simple linear regression model fit. The asterisk indicates a p < .05 significance level.

The severity of negative symptoms further positively correlated with the connectivity strength from the LHC to the LIPAR (*r* = .41), and a simple linear regression model (see Figure 6. B) additionally, showed a significant relation between the two variables (*p* = .05). The slope coefficient was 0.046, and the adjusted R^2^ value was 0.1312.

The severity of negative symptoms was also negatively correlated to the connectivity strength from the LHC to the RHC (*r* = −.41), and a simple linear regression model (see Figure 6. C) additionally, showed a significant relation between the two variables (*p* = .05). The model’s slope coefficient was − 0.028, and the adjusted R^2^ value was 0.131.

#### Positive Symptoms Severity and Connectivity Strength

The severity of positive symptoms was negatively correlated to the connectivity strength from the RHC to the LHC (*r* = − .51). A simple linear regression model (see Figure 7. A) further showed that the relation between these variables was significant (*p* = .013), with a slope of – 0.05 and an adjusted R^2^ value of 0.2242.

**Figure 7.**
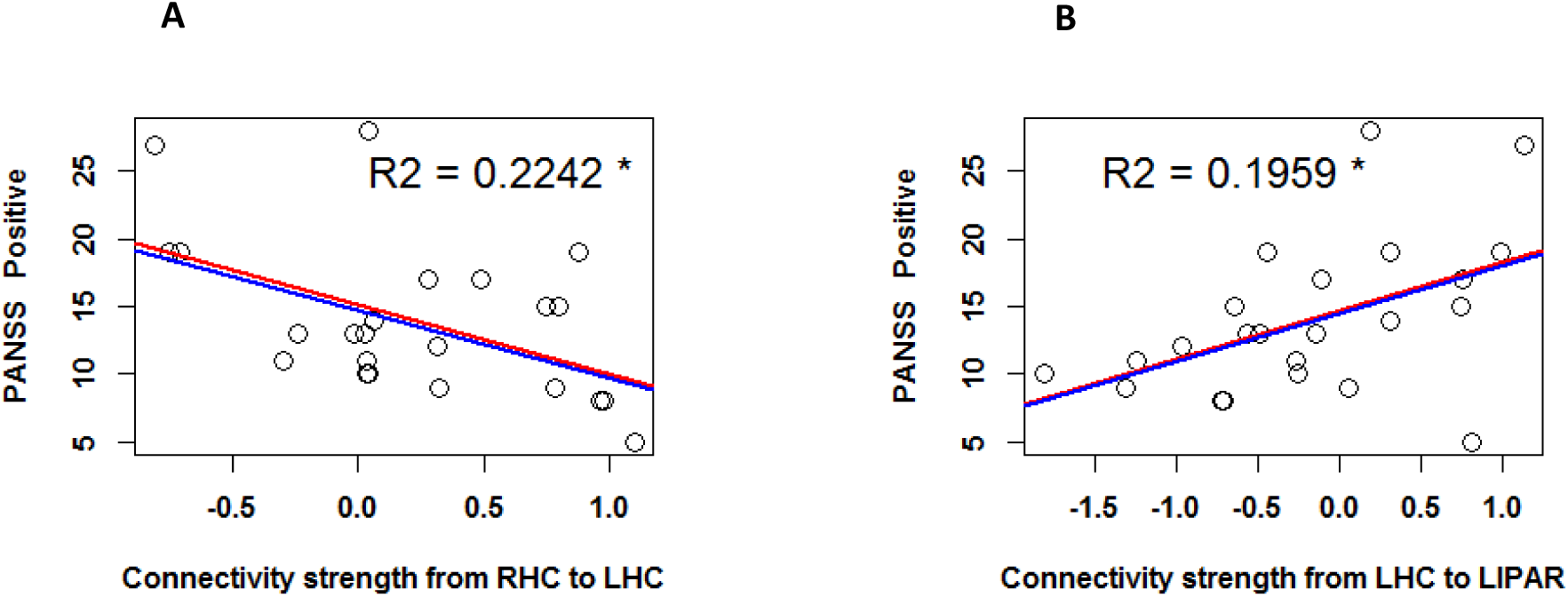
Linear regression with severity of positive symptoms (as reflected by PANSS scores) as regressor. The predicted variables are the connectivity strengths of the following parameters **A**: from the RHC to the LHC, and **B**: from the LHC to the LIPAR. The red regression line reflects the fit of the simple linear regression model, while the blue line reflects that of the robust regression model. The plotted R^2^ values reflect the simple linear regression model fit. The asterisk indicates a p < .05 significance level.

Finally, the severity of positive symptoms was positively correlated to the connectivity strength from the LHC to the LIPAR (*r* = .48). A simple linear regression model (see Figure 7. B) further showed that the relation between these variables was significant (*p* = .009), with a slope of 0.07 and an adjusted R^2^ value of 0.1959.

### 3.3. Voxel based morphometry (VBM) Analysis of Gray Matter Volume (GMV)

We analyzed group differences of GMV for all the 13 nodes of our RSNs using an ANCOVA with total intracranial volume (TIV) as covariate. After controlling for TIV, a main effect of group remained significant for the following nodes: MPFC, LIPAR, DACC, LAI, RAI, LFEF, RFEF, LIPS. Post-hoc comparisons using Welch t-test revealed that HC had significantly higher GMV than SZ in the following nodes: MPFC, LIPAR, DACC, LAI, RAI, RFEF, LIPS. These results, as well as effects sizes are reported in Table 3 below.

**Table 3:** Mean gray matter volume (GMV) of each region of interest (ROI) and mean total intra-cranial volume (TIV) per group. Group differences in GMV were analyzed with ANCOVA using TIV as covariate. The results of post-hoc analyses with Welch two samples t-tests, as well as effect sizes are also summarized.

| ROI | GMV Mean (SD) |  | HC vs. SZ (adjusted for TIV) |  |  | HC > SZ |  |  |  |
| --- | --- | --- | --- | --- | --- | --- | --- | --- | --- |
| | HC | SZ | F(1) | *p* | $\eta_p^2$ | *t* | df | *p* | *d* |
| MPFC | 0.53 (0.03) | 0.49 (0.05) | 10.86 | .002 | .17 | 3.15 | 41 | .003 | 0.88 |
| LIPAR | 0.49 (0.03) | 0.46 (0.05) | 6.61 | .013 | .11 | 3.15 | 39 | .014 | 0.72 |
| RIPAR | 0.50 (0.04) | 0.49 (0.05) | 0.53 | .47 | .01 | 0.65 | 47 | .52 | 0.18 |
| PCC | 0.60 (0.06) | 0.59 (0.06) | 1.1 | .30 | .02 | 0.61 | 48 | .54 | 0.17 |
| LHC | 0.54 (0.04) | 0.52 (0.04) | 2.92 | .09 | .05 | 1.79 | 51 | .08 | 0.48 |
| RHC | 0.54 (0.03) | 0.53 (0.04) | 2.94 | .09 | .05 | 1.14 | 47 | .26 | 0.31 |
| DACC | 0.52 (0.04) | 0.48 (0.04) | 18.33 | <.001 | .26 | 3.57 | 50 | <.001 | 0.97 |
| LAI | 0.56 (0.05) | 0.53 (0.05) | 10.16 | .002 | .16 | 2.23 | 47 | .03 | 0.61 |
| RAI | 0.60 (0.05) | 0.56 (0.05) | 12.25 | <.001 | .19 | 2.80 | 51 | .007 | 0.75 |
| LFEF | 0.53 (0.07) | 0.50 (0.05) | 5.1 | .03 | .10 | 1.74 | 53 | .09 | 0.45 |
| RFEF | 0.51 (0.06) | 0.47 (0.05) | 14.18 | <.001 | .21 | 2.95 | 54 | .005 | 0.77 |
| LIPS | 0.49 (0.07) | 0.45 (0.04) | 6.42 | .014 | .11 | 2.73 | 50 | .009 | 0.72 |
| RIPS | 0.49 (0.06) | 0.47 (0.05) | 2.81 | .10 | .05 | 1.53 | 54 | .13 | 0.40 |
| TIV | 1.59 (0.11) | 1.54 (0.14) |  |  |  | 1.51 | 44 | .14 | 0.42 |

#### GMV and Connectivity Strength

We additionally ran an ANCOVA analysis to check whether group differences in directed connectivity strengths remained significant when accounting for the GMV of both RSNs nodes forming each pair of directed connectivity. Group differences in directed connectivity that remained significant even after controlling for GMV group differences were the following: PCC −> MPFC, PCC −> DACC, LHC −> RHC, LHC −> LIPAR, RFEF −> LHC, LIPS −> RHC, RIPS −> DACC (see Table 4 below).

**Table 4:** Group differences in mean directed connectivity strengths for each region of interest (ROI). The ANCOVA with GMV of each ROI as covariate, Welch two samples t-tests and effect sizes are also summarized.

| Connection | Connectivity strength<br>Mean (SD) |  | HC vs. SZ<br>(adjusted for GMV of both nodes) |  |  | HC > SZ |  |  |  |
| --- | --- | --- | --- | --- | --- | --- | --- | --- | --- |
| | HC | SZ | F(1) | P | $\eta_p^2$ | t | df | p | d |
| MPFC -> LHC | 0.05<br>(0.33) | 0.24<br>(0.47) | 3.54 | .07 | .06 | -1.75 | 41 | .08 | -0.49 |
| PCC -> RAI | 0.03<br>(0.37) | -0.16<br>(0.47) | 3.87 | .06 | .07 | 1.94 | 45 | .06 | 0.54 |
| PCC -> MPFC | 0.03<br>(0.43) | -0.18<br>(0.34) | 4.24 | .05 | .08 | 2.08 | 54 | .05 | 0.54 |
| PCC -> RHC | 0.04<br>(0.44) | -0.19<br>(0.41) | 3.47 | .07 | .06 | 1.96 | 53 | .06 | 0.52 |
| PCC -> DACC | 0.14<br>(0.3) | -0.11<br>(0.4) | 7.3 | .01 | .12 | 2.58 | 43 | .01 | 0.72 |
| LHC -> RHC | 0.5<br>(0.46) | 0.18<br>(0.51) | 5.01 | .03 | .09 | 2.11 | 49 | .04 | 0.57 |
| LHC -> LIPS | -0.12<br>(0.78) | 0.09<br>(0.81) | 0.79 | .38 | .02 | -0.99 | 51 | .33 | -0.27 |
| LHC -> LIPAR | 0.17<br>(0.6) | -0.25<br>(0.8) | 5.3 | .03 | .09 | 2.16 | 44 | .04 | 0.6 |
| LHC -> DACC | -0.27<br>(0.44) | -0.06<br>(0.74) | 0.99 | .32 | .19 | -1.27 | 37 | .21 | -0.36 |
| LHC -> RIPS | -0.001<br>(0.57) | 0.27<br>(0.7) | 2.64 | .11 | .05 | -1.59 | 46 | .12 | -0.44 |
| LHC -> RIPAR | 0.28<br>(0.79) | -0.04 | 1.9 | .17 | .04 | 1.59 | 54 | .12 | 0.42 |

|  |  | (0.05) |  |  |  |  |  |  |  |
| --- | --- | --- | --- | --- | --- | --- | --- | --- | --- |
| LHC -> RAI | -0.44<br>(0.54) | 0.33<br>(0.9) | 3.11 | .08 | .06 | -1.84 | 38 | .08 | -0.52 |
| LHC -> PCC | 0.45<br>(0.66) | 0.22<br>(1.03) | 0.88 | .35 | .02 | 0.95 | 39 | .35 | 0.27 |
| LHC -> LFEF | -0.03<br>(0.56) | 0.2<br>(0.53) | 2.76 | .10 | .05 | -1.6 | 53 | .12 | -0.43 |
| RHC -> RAI | -0.13<br>(0.85) | 0.08<br>(0.94) | 1.04 | .31 | .00 | -0.87 | 49 | .39 | -0.24 |
| RHC -> RFEF | 0.08<br>(0.55) | 0.24<br>(0.44) | 2.64 | .11 | .01 | -1.23 | 54 | .23 | -0.33 |
| RFEF -> LHC | 0.14<br>(0.38) | -0.05<br>(0.33) | 3.57 | .07 | .07 | 2.03 | 54 | .05 | 0.54 |
| RFEF -> RIPS | 0.33<br>(0.41) | 0.11<br>(0.47) | 2.68 | .11 | .05 | 1.91 | 49 | .06 | 0.52 |
| RHC -> LHC | 0.45<br>(0.5) | 0.22<br>(0.55) | 3.55 | .07 | .06 | 1.61 | 50 | .12 | 0.44 |
| RHC -> RIPS | -0.03<br>(0.93) | 0.3<br>(0.64) | 2.03 | .16 | .04 | -1.53 | 53 | .13 | -0.4 |
| RHC -> LAI | -0.3<br>(1.21) | 0.08<br>(0.81) | 2.16 | .15 | .04 | -1.41 | 52 | .17 | -0.36 |
| LIPS -> RHC | 0.07<br>(0.38) | -0.18<br>(0.37) | 5.57 | .03 | .97 | 2.44 | 52 | .02 | 0.66 |
| RIPS -> DACC | 0.04<br>(0.33) | -0.19<br>(0.41) | 5.2 | .03 | .09 | 2.23 | 46 | .03 | 0.61 |
| RAI -> LHC | 0.16<br>(0.4) | -0.02<br>(0.36) | 4.03 | .05 | .07 | 1.82 | 53 | .08 | 0.48 |

## 4. Discussion

The dysconnection hypothesis posits that schizophrenia symptoms arise from impaired brain network function, and not (only) from discrete structural and/or functional alterations (Friston & Frith, 1995). Since a network is characterized by directed interactions between brain areas, effective connectivity methods, especially DCM, appear to be preferable to the undirected (i.e. functional) ones. These directed approaches have demonstrated increased reliability (e.g. Schuyler et al., 2010) and can point to potential *intermediate phenotypes* (i.e. markers of heritability) in the study of schizophrenia (Cao et al., 2016).

In this study, we investigated group differences in effective connectivity between HC and SZ with respect to brain areas which are known to play a role in three major RSNs: DMN, SAN and DAN. Of particular interest to us were the bilateral hippocampi (i.e. LHC and RHC), which have been found to show distinctive connectivity patterns in SZ, as well as volumetric alterations. We sought to explore a potential link between GMV and directed connectivity strengths by checking whether the strength of directed connectivity patterns in SZ persisted when controlling for GMV of the node pairs. Additionally, we also wanted to see whether symptom severity could predict connection strengths. Finally, we wanted to assess whether hierarchical strengths of and between the three RSNs revealed any group differences. To our knowledge, this is the first paper which investigated hierarchical strength differences of large scale RSNs between SZ and HC. Additionally, even though these three networks have been previously investigated from a directed connectivity perspective in SZ (e.g. Zhou et al., 2018b), it has been done so during a task-based memory paradigm and not at rest. We have however chosen to investigate group differences in directed connectivity between these three networks during rest. Given the practicality of this acquisition modality, we are confident that future studies involving even more severely affected patients could more easily report relatable results.

Our current results largely speak in favour of the dysconnection hypothesis in SZ. We found no less than 24 distinct patterns of directed connectivity which were significantly different between HC and SZ, most of these originating in the PCC, LHC and RHC (nodes belonging to the DMN). In SZ, the connections from the PCC to other DMN and SAN nodes were significantly weaker in comparison to HC, but only the PCC to DACC connection strength group difference remained significant when controlling for GMV of both nodes. Additionally, the connections from the LHC to other DMN nodes were significantly weaker in SZ than in HC, while the connections from the LHC to the SAN and DAN nodes were significantly stronger in SZ than in HC. Finally, the connections from the LHC to other nodes of the SAN and DAN were also significantly stronger than in HC, but weaker towards other DMN nodes. After controlling for the GMV of both node pairs however, only the connection strengths from the LHC to RHC and to LIPAR remained significantly higher in HC. Overall, these results point to a general pattern of exacerbated connectivity strength from the nodes of the DMN to those of the SAN and DAN in the SZ group.

Previously, Zhou et al. (2018a) have revealed that a hierarchical relation between the nodes of these three networks occurs in the healthy general population, with nodes of the DMN exciting those of the SAN and the DAN. Our results therefore suggest that even though this hierarchy is maintained in SZ, the strength of connections within this hierarchy are significantly increased compared to what can be seen in HC. As opposed to Zhou et al. (2018a) however, we were unable to replicate this hierarchical relationship in our HC group; on the contrary, in our control sample, the DMN exited both the DAN and the DAN. One important distinction between their study and ours which might have led to this divergent finding is that Zhou et al. considered the core DMN only, without including the hippocampi. For our study, however, given our special interest in hippocampal dysconnectivity in SZ, we consider that an extended DMN conceptualization is highly relevant. Additionally, these authors included additional areas in their SAN (i.e. bilateral anterior prefrontal cortices) and DAN (i.e. bilateral inferior frontal gyri). A complete replication of their results might therefore be dependent on the initial definition of the respective RSNs.

An additional indication that the hippocampi might play a special role in SZ dysconnectivity was our finding that the strength of directed connectivity originating in the hippocampi were modulated by the severity of negative and positive symptoms. Specifically, the connection strength from the LHC to the LIPAR was positively predicted by the intensity of both positive and negative symptoms. Additionally, the connection strengths from the LHC to the DACC and from the LHC to the RHC were negatively predicted by the intensity of negative symptoms. Finally, the intensity of positive symptoms negatively predicted the connection strength from the RHC to the LHC. We therefore show that hippocampal connections to nodes of the SAN (such as the DACC) can be predicted by the severity of negative symptoms. In addition to previous research (e.g. Duan et al., 2015; Garrity et al., 2007) which found exclusively predictive relationship between connectivity strengths and positive symptom severity, we found that negative symptom severity was also a significant predictor. A potential explanation for this may reside in our choice of a DCM based directed connectivity assessment, i.e. relating symptom severity to the strength of directed connections between two nodes rather than to the time courses of one single node.

In conclusion, our results support a robust relation between dysconnectivity, symptom severity and volumetric alterations in in SZ. Moreover, we trust that our approach, based on exploring directed dysconnectivity characteristics of large-scale RSNs and their relation to symptoms and structure can offer valuable insight into putative mechanisms that give rise to this disorder.

The main limitation of the current study resides in its rather small sample size. A suitable follow-up aimed at overcoming this drawback would therefore be the replication of our directed connectivity analyses on an independent, larger sample, such as The Center for Biomedical Research Excellence (COBRE; http://fcon_1000.projects.nitrc.org/indi/retro/cobre.html). Finally, while all our patients were medicated and stable, we were unable to control for the potential effect that their medication might have had on their connectivity strengths or their GMV. With respect to GMV, previous studies have shown that medication might act to attenuate the differences between SZ and HC (e.g. Ho et al., 2011), so a longitudinal assessment of pre- and post-medication SZ might offer a more accurate view on GMV group differences. Especially with respect to the hippocampus, although we found no significant GMV group differences in our sample, previous authors (e.g. Radulescu et al., 2014) have reported reduced volumes in SZ compared to HC. Nevertheless, total hippocampal GMV group differences have not always been replicated, which led some authors to recommend a sub-hippocampal structures approach instead (e.g. Folley et al., 2010), or multimodal methodologies (e.g. Boyer et al., 2007).

## Data Availability

All data is available upon request.

## Contributors

Author 10 designed the study and wrote the protocol. Author 2 wrote the acquisition parameters section. Authors 3,4,5,6,7 provided access to patients and aided in their assessment. Authors 1 and 10 analyzed the data. Author 1 wrote the manuscript. All authors contributed to the final version of the submitted manuscript.

## Acknowledgements

We wish to thank Peter Zeidmann for his valuable advice regarding our application of spDCM. Additionally, we wish to thank Derek Evan Nee for his advice on how to compute the network hierarchical strength.

